# Voxel-wise temporal decomposition of hypoxia-powered BOLD MRI: method development and proof-of-concept application in glioblastoma

**DOI:** 10.64898/2026.05.27.26354265

**Authors:** Tristan Schmidlechner, Vittorio Stumpo, Elisabeth Jehli, Leonie Zerweck, Jacopo Bellomo, Meltem Gönel, Flavia Müller, Natalia Cantavella Franch, Martina Sebök, Andrea Bink, Zsolt Kulcsár, Michael Weller, Luca Regli, Jorn Fierstra, Christiaan H. B. van Niftrik

## Abstract

Hypoxia-powered blood oxygen level-dependent (BOLD) MRI probes the BOLD response to a controlled transient hypoxic stimulus. We developed GLIODISSECT, an operational voxel-wise framework that separates relative response arrival, descending and recovery transitions, and threshold-defined steady state amplitude. Twenty healthy controls underwent 3-T BOLD MRI during a double hypoxic-step challenge, and three patients with newly diagnosed glioblastoma were included as proof-of-concept cases. Full-record cross-correlation and first-step 10% landmarks yielded one refined voxel-wise lag that was reused downstream. Alternative 5%/95% and 20%/80% thresholds preserved steady state map patterns (median within-subject spatial r 0.9987-0.9998), while timing measures were more threshold-dependent. Numerical criteria were satisfied in 97.789% and 97.682% of evaluable delay-to-plateau (DTP) estimates and in all evaluable delay-to-return (DTR) estimates. Leave-one-subject-out validation of the global-response-adjusted steady state model yielded a median voxel-wise Q^2^ of 0.345 and reduced pooled prediction error by 16.0%, improving 19 of 20 controls. In the three illustrative cases, component and deviation maps displayed spatially heterogeneous timing and steady state patterns that could not be represented by one response-amplitude summary. GLIODISSECT supports component-specific mapping and voxel-wise normative interpretation. Biological and clinical validation remain required.

## 1 Introduction

Glioblastoma is the most common malignant primary brain tumor [1]. Its infiltrative component extends beyond the contrast-enhancing border and is difficult to distinguish from surrounding edema on conventional MRI [2, 3, 4]. Delineating this tissue is important for planning the extent of resection [5]. Vascular dysregulation has been reported in non-enhancing peritumoral tissue [6, 7]. Studies of diffuse glioma also demonstrate cerebrovascular reactivity (CVR) abnormalities beyond MRI-defined tumor margins and in whole-brain measures [8, 9, 10]. Hypoxia-powered blood oxygen level-dependent (BOLD) MRI has been applied to glioblastoma using controlled oxygen challenges to map spatially heterogeneous tissue responses [11, 12]. These responses vary across voxels in both timing and magnitude. A single response-amplitude summary cannot distinguish delayed arrival, prolonged transition, steady state, and recovery. The present work adapts the established temporal decomposition framework from CVR imaging to hypoxia-modulated BOLD MRI [13]. A recently published narrative review summarizes the physiological principles, technical variability, and emerging glioblastoma applications of hypoxia-modulated BOLD MRI [14]. Here, temporal decomposition denotes sequential threshold-based parameterization of a voxel time course into operational timing intervals and a steady state amplitude estimate. It is not an additive signal decomposition, and the derived metrics are neither statistically independent nor exhaustive. We aimed to establish and technically validate the method, construct a voxel-wise normative reference, and demonstrate its interpretive potential in three proof-of-concept glioblastoma cases. We did not aim to establish diagnostic superiority over an existing benchmark, because no validated reference analysis exists for this double-hypoxia paradigm.

## 2 Materials and Methods

### 2.1 Ethics Statement

The study was approved by the Cantonal Ethics Committee Zurich (KEK-ZH ID 2020-02314, approval date: November 17th, 2020). All procedures were conducted in accordance with the Declaration of Helsinki and applicable institutional and national regulations. Written informed consent was obtained from all participants before MRI acquisition. The privacy rights of all participants were observed.

### 2.2 Study Overview and Analysis Framework

This study enrolled 20 healthy subjects (12 female, 8 male, age 27 ± 4 years, range 22-40 years) between October 22, 2022 and February 2, 2026. Voxel-wise maps were generated for four operational features: Delay_corr_ = (i_10,voxel_ - i_10,reference_) × repetition time (TR), delay to plateau (DTP) = (i_90,descent_ - i_10,descent_) × TR, delay to return (DTR) = (i_10,recovery_ - i_90,recovery_) × TR, and Hypoxia_SS_, the mean baseline-referenced BOLD change in the threshold-defined steady state window divided by the achieved change in end-tidal oxygen partial pressure (ΔP_ET_O_2_). The first-step threshold landmarks also refined one voxel-wise total lag used internally for downstream window placement. This alignment quantity is distinct from the reported relative Delay_corr_ map. Detailed definitions are provided below. A voxel-wise normative atlas was constructed from the healthy controls in Montreal Neurological Institute (MNI) space and used to characterize normal spatial response patterns. Patient parametric maps were retained in native space, and the normative atlas was registered to each patient to avoid spatial distortion from tumor-related mass effect.

### 2.3 Controlled Transient Hypoxia Stimulus

All subjects underwent the same controlled transient double-hypoxia stimulus using a computer-controlled prospective gas-targeting system (RespirAct Gen4, Thornhill Medical, Toronto, Canada). Prospective gas targeting permits independent control of end-tidal oxygen and carbon dioxide [15, 16]. Here, end-tidal oxygen partial pressure (P_ET_O_2_) was targeted while end-tidal carbon dioxide was held constant (isocapnia). The stimulus comprised two 60-second hypoxic steps targeting P_ET_O_2_ 40 mmHg, separated by a 40-second normoxic interval. Because the physiological response is sampled only at the BOLD repetition time and remains affected by noise and regional transit, the protocol does not provide direct measurements of tissue oxygen tension, blood flow, or oxygen metabolism.

### 2.4 MRI Data Acquisition

All subjects were examined on a 3-Tesla Skyra VE11 scanner (Siemens, Erlangen, Germany) using a 32-channel head coil. BOLD MRI data were planned on the anterior commissure–posterior commissure (AC-PC) line plus 20° anticlockwise on a sagittal localizer and acquired using T2*-weighted gradient-echo echo-planar imaging (EPI) with the following parameters: repetition time (TR) = 1800 ms, echo time (TE) = 30 ms, slice thickness = 2.5 mm, 50 slices acquired in interleaved order, field of view (FOV) = 220 *×* 220 mm^2^, flip angle (FA) = 80°, 160 measurements, acquisition time (TA) = 5:00 min.

### 2.5 Data Preprocessing

All MRI data were preprocessed in SPM12 (Wellcome Centre for Human Neuroimaging, London, UK), followed by custom MATLAB R2025b Update 4 routines adapted from the published CVR temporal decomposition implementation [13]. Functional BOLD images underwent slice-timing correction, rigid-body realignment, co-registration to the subject’s T1 image, and spatial smoothing with a 6-mm isotropic Gaussian kernel. For anatomical localization in the patient cases, T1 post-contrast, T2, and fluid-attenuated inversion recovery (FLAIR) images were co-registered to the native T1 image and normalized with the same deformation field. A brain analysis mask was generated from the available tissue and lesion masks. The patient analysis domain included tissue-or-lesion support and was not restricted to pure grey and white matter. The masked BOLD time series was linearly detrended with mean restoration, low-pass filtered at 0.125 Hz, and smoothed using centered robust locally estimated scatterplot smoothing (Loess, 12-volume window, 21.6 s). These fixed preprocessing settings were applied before lag and threshold estimation.

### 2.6 Gas Alignment and Voxel-wise Oxygen Reference Trace

The recorded P_ET_O_2_ waveform was resampled to the BOLD temporal grid using the piecewise cubic Hermite interpolating polynomial (PCHIP) method. PCHIP preserves local shape and monotonic transitions while avoiding spline overshoot. Resampling does not create physiological timing information beyond the original gas sampling and the 1.8-s BOLD repetition time. Initial global and voxel-wise lags were estimated over the full recording by cross-correlation of the detrended, normalized BOLD and oxygen traces over actual lag labels 0-15 TR. Absolute correlation was used, with the first maximum selected in a tie. These initial estimates served only to initialize first-step landmark detection. After first-step refinement, the rounded fifth percentile of finite in-mask refined total lags aligned the global oxygen trace for subsequent calculations.

### 2.7 Temporal Decomposition

#### 2.7.1 Iterative Estimation of Voxel-wise Arrival Delay (Delay_corr_)

Voxel-wise timing was initialized from the full-record cross-correlation lag and refined from the first hypoxic step. DTP1 was calculated once with the initial voxel-wise lag. Its retained 10% response index (i_10_), the programmed first-step timing, and the initial global lag then defined a refined voxel-wise total lag. DTP1 was not recalculated after refinement. The refined first-step total lag was reused for DTR1, DTP2, DTR2, both steady state maps, and both whole-stimulus averages. No separate step-2 lag was calculated. Because the initializer uses the complete recording and the established search windows can extend beyond the nominal step boundary, the procedure should not be interpreted as sample-level independence of step 1 from later time points. Mean baseline (S_baseline_) and step (S_step_) signals defined voxel-specific 10% and 90% levels from the baseline-to-step excursion (Equations 1a and 1b).

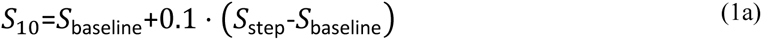

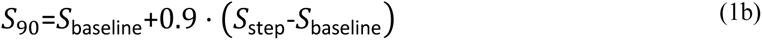

Because the hypoxic challenge produces a signal decrease, threshold crossings were evaluated according to the sign of the voxel-specific response. An initial i_90_ was identified as the first admissible 90% crossing after stimulus onset, and i_10_ was identified by backward search from i_90_ to the first 10% crossing. These provisional indices redefined the effective baseline and response windows. The means, thresholds and crossings were then recalculated. For DTP, two unconditional refinements were followed by up to 15 conditional refinements, permitting a maximum of 17 completed refinements. During conditional stages, successive baseline and step means were compared with relative-change bounds of 0.9999-1.0001. If the bound was reached, the final bounded estimate was retained. Prespecified finite crossing fallbacks could be encountered during the search. Fallback encounter, criterion satisfaction and validity of the final output were recorded separately. Failed calculations or invalid final indices were retained as missing values. Figure 1 provides a schematic overview.

**Fig. 1:**
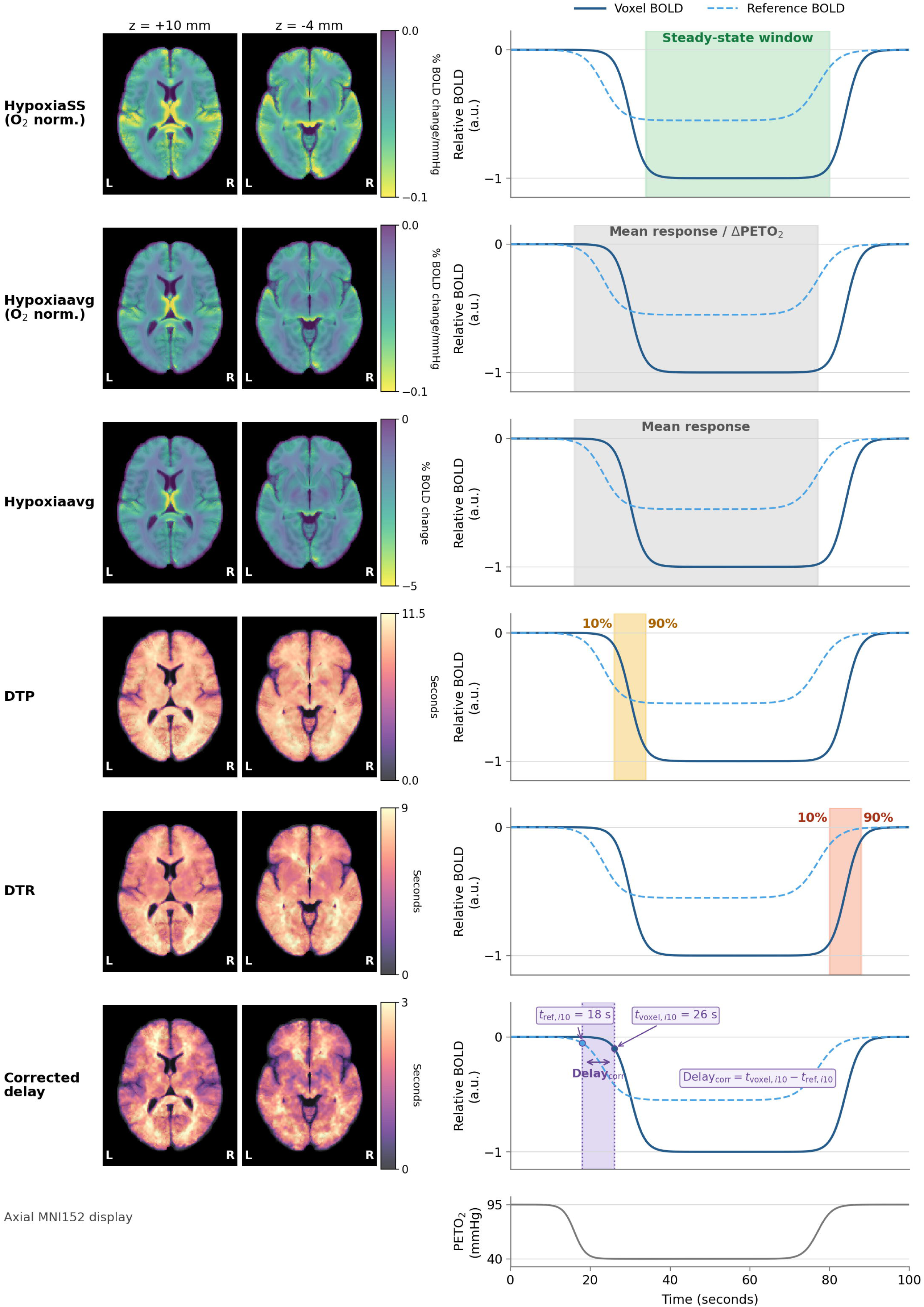
Hypoxia-powered BOLD response metrics and their temporal definitions. Left: voxel-wise mean maps from the 20-control normative atlas at MNI axial levels z = +10 mm and z = −4 mm. Response maps use reversed viridis (oxygen-normalized responses −0.10 to 0 %BOLD/mmHg, raw average −5 to 0 %BOLD), and timing maps use magma (DTP 0-11.5 s, DTR 0-9 s, relative arrival delay 0-3 s). The row labelled corrected delay displays the reporting quantity Delay_corr_, not the refined total lag used for alignment. Right: illustrative time courses with relative BOLD amplitude in arbitrary units and time in seconds. Solid and dashed curves illustrate a voxel and an earlier reference response. Their amplitudes are schematic, not measured %BOLD. Green shading marks the steady state window, grey shading the whole-stimulus interval, orange shading the descending 10%-90% transition, and red shading the recovery transition. The DTR panel labels recovery progress from 10% to 90%, equivalent to moving from the 90% to the 10% level of the baseline-to-step excursion. The purple interval illustrates Delay_corr_ = t_voxel,i10_ - t_ref,i10_ (26 - 18 = 8 s in the schematic). In the analysis, t_ref,i10_ is the unrounded within-subject 5th-percentile arrival time and need not correspond to an individual voxel. The bottom panel shows a schematic end-tidal oxygen trace. These diagrams explain the metrics. Numerical window placement follows the Methods.

The final retained first-step i_10_ served two distinct purposes. First, together with the initial lag and timing origin, it refined the voxel-wise total lag used for downstream window placement. Second, a reporting-only within-subject arrival reference was defined as the unrounded 5th percentile of valid voxel-wise i_10_ values. Delay_corr_ is the voxel i_10_ minus this reference (Equation 2), so negative and zero relative values are valid. The rounded fifth percentile of refined total lag used for global oxygen alignment is a different quantity. DTP and DTR share threshold landmarks with Delay_corr_, whereas Hypoxia_SS_ uses i_90_ to define its steady state window. The outputs are therefore mathematically related and may be correlated. They are not independent, additive, or exhaustive components of the BOLD signal.

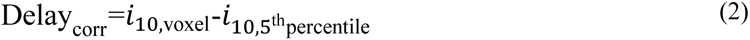

Delay_corr_, DTP and DTR are computed in scan intervals by Equations 2-4 and converted to seconds for reporting and display by multiplying by TR (1.8 s). Delay_corr_ is relative to the within-subject 5th-percentile i_10_ reference. It is distinct from the refined total lag used internally for alignment.

#### 2.7.2 Transition Timing (DTP and DTR)

Delay to plateau (DTP) was defined as the interval between the final retained 10% and 90% threshold crossings on the descending hypoxic response (Equation 3):

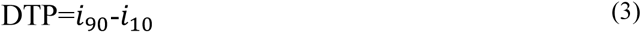

Delay to return (DTR) was defined as the interval between the 90% and 10% threshold crossings during recovery (Equation 4):

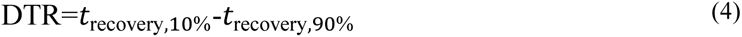

DTR used a separate iterative recovery procedure with up to 15 refinements and stopped when the absolute relative change in successive baseline means was below 0.0001. Both metrics were computed for each hypoxic step.

### 2.8 Parametric Maps

#### 2.8.1 Steady-State Response (Hypoxia_SS_)

Hypoxia_SS_ was defined operationally as the mean %BOLD signal change during the threshold-defined steady state window of each hypoxic step, excluding the descending and recovery transition intervals. The window began at the 90% signal descent threshold and ended at the earlier of the 90% recovery crossing or the end of the refined-lag-adjusted oxygen stimulus reference (Equation 5). Step 2 reused the first-step refined lag and the first-step baseline definition. Here, steady state denotes the threshold-defined BOLD plateau and does not by itself establish physiological equilibration.

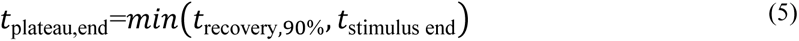

Because the endpoint is shifted by the refined voxel-wise total lag, the cap does not apply the same uncorrected physical time to every voxel. It prevents extension of the averaging window beyond the hypoxic portion of that voxel’s aligned oxygen reference. Each subject’s steady state map was normalized by the achieved end-tidal oxygen step amplitude (ΔP_ET_O_2_) (Equation 6).

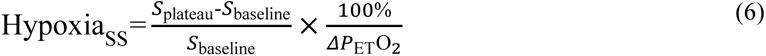

where S_plateau_ is the mean BOLD signal during the threshold-defined steady state window and S_baseline_ is the first-step normoxic baseline mean, reused for both steps. Hypoxia_SS_ is reported in %BOLD/mmHg. It is a scale-adjusted response metric rather than a fitted regression slope and should not be interpreted as a direct measurement of tissue oxygenation, blood flow, oxygen extraction, or metabolic equilibration.

#### 2.8.2 Whole-Stimulus Reference Map

For visual context in the proof-of-concept patient figures, a whole-stimulus reference map (Hypoxia_avg_) was computed with the same refined voxel-wise total lag as Hypoxia_SS_. For each step, the full lag-adjusted stimulus block was averaged and referenced to the equally weighted mean of the three normoxic baseline-period means (S_baseline,avg_, Equation 7):

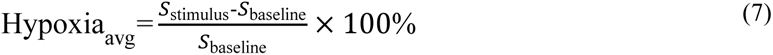

A scale-matched visual reference was then computed by dividing Hypoxia_avg_ by the achieved ΔP_ET_O_2_ (Equation 8):

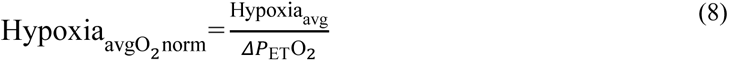

The oxygen-normalized whole-stimulus map was retained only as a simple visual summary of the complete stimulus period. It includes the descending transition and steady state periods and is therefore not a biological benchmark for Hypoxia_SS_. No formal superiority analysis was based on this reference map.

### 2.9 Tumor Segmentation

Tumor sub-compartment masks were derived from the multimodal structural images using the automated Oncohabitats framework [17], yielding contrast-enhancing tumor, necrosis and FLAIR abnormality masks. These masks served two purposes: to exclude tumor tissue from the brain parenchyma mask used for global response scalar (G) computation and to provide anatomical localization relative to the structurally defined tumor compartments.

### 2.10 Normative Atlas Construction and Deviation Mapping

Normative modelling characterizes individual deviations relative to a reference population [18]. A voxel-wise normative atlas was constructed from the 20 healthy controls. Conventional references used each map’s mean and sample standard deviation in MNI space, with finite-map coverage in at least 16 controls. The standard-deviation floor was the 5th percentile of positive finite values within that map’s eligible domain, applied before resampling to patient space. For Hypoxia_SS_, a G-adjusted model was fitted by voxel-wise ordinary least squares. Each control’s G was the median finite native-T1 oxygen-normalized step-1 Hypoxia_SS_ response within the prespecified individual G mask. Model eligibility required individual registered grey matter (GM) + white matter (WM) probability of at least 0.8 and finite response coverage in at least 16 controls. In-sample R^2^ and leave-one-subject-out predictive Q^2^ were reported separately. In each of 20 folds, only the 19 training controls defined anatomical and response eligibility (at least 16 training controls), model coefficients and the residual-SD floor. The held-out control was compared with the G-adjusted prediction and the mean of the same valid training observations. Voxel-wise Q^2^ required at least 17 scored held-out predictions. Pooled root-mean-square error (RMSE) combined all scored held-out errors. Patient G_p_ used tumor-excluded support, which differs from control support and may influence the patient-specific reference. Reference maps were resampled to native patient T1 space using the inverse SPM12 deformation, with fourth-order interpolation for continuous maps and nearest-neighbour interpolation for support. G-adjusted residual SD was floored after resampling. Conventional SD retained its pre-resampling floor and was required to remain finite and positive afterward. No z-score clipping was applied. Hypoxia_SS_ deviation maps used G-adjusted standardized residuals. Whole-average and timing maps used conventional z-scores (Equation 9).

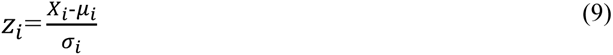

For conventional z-scores, X_i_ is the patient value at voxel i and μ_i_ and σ_i_ are the atlas mean and sample standard deviation at that voxel. For G-adjusted Hypoxia_SS_, the corresponding reference is β_0,i_ + β_1,i_G_p_ and the denominator is the model residual standard deviation. Voxels with |z| > 2 were displayed descriptively as deviations from the healthy reference. Structural lesion masks were used for anatomical orientation only and did not provide biological ground truth.

### 2.11 Statistical Analysis

Analysis was performed with MATLAB R2025b Update 4. The primary analyses evaluated the numerical behavior of the temporal decomposition, sensitivity to alternative transition thresholds, and out-of-sample performance of the global-response-adjusted normative model. The three patient cases were intentionally descriptive and no inferential or group-level patient statistics were performed. The whole-stimulus maps were displayed as visual context only. They were not treated as a validated benchmark or used to rank the methods.

### 2.12 Sensitivity Analysis

Parameter sensitivity was evaluated in all 20 healthy controls. The reference configuration used 10%/90% transition thresholds, 6-mm spatial smoothing and a 12-volume robust-Loess temporal window. Each alternative changed one factor only: thresholds of 5%/95% or 20%/80%, temporal windows of 8 or 16 volumes, or spatial smoothing of 4 or 8 mm. For each subject and decomposition output, the alternative native-space map was compared with the reference over common finite support using spatial Pearson correlation, median signed and absolute differences, and response-magnitude change. Coverage was recorded separately. Smoothing analyses also recorded root-mean-square error. Group summaries are medians and interquartile ranges across subjects.

Response and timing quantities were evaluated separately because changing a transition threshold changes the operational definition of the timing intervals. High spatial correlation was not interpreted as unchanged magnitude, calibration or patient abnormality classification.

### 2.13 Iteration-count and Termination Analysis

Iteration and termination behavior was evaluated in all 20 healthy controls using instrumented versions of the analysis functions that preserved numerical outputs. Refinement counts excluded initialization. Criterion satisfaction, reaching the iteration bound, crossing or window fallback encounters, caught errors, and invalid final outputs were recorded separately. DTP used two unconditional plus at most 15 conditional refinements. DTR used at most 15 refinements. Regional and pooled summaries were descriptive and did not assume voxel independence.

## 3 Results

### 3.1 Normative Atlas

A voxel-wise normative atlas was constructed from 20 healthy controls. After MNI normalization and the predefined 16-of-20 coverage requirement, mean maps showed consistent spatial organization across the plateau and temporal metrics (Figure 1). Group-level healthy-control reference and normative-model maps are available on Mendeley Data: https://doi.org/10.17632/2k4xh888b4.1.

### 3.2 Sensitivity Analysis

All six alternative parameter configurations completed successfully in all 20 controls. Relative to 10%/90%, steady state map patterns were highly preserved, with median within-subject spatial r ranging from 0.9987 to 0.9998 and median response-magnitude changes from −1.57% to +0.69%. Changing the robust-Loess temporal window from 12 volumes to 8 or 16 volumes yielded Hypoxia_SS_ median spatial r of 0.9975-0.9991. Changing spatial smoothing from 6 mm to 4 or 8 mm had a larger effect (Hypoxia_SS_ median r 0.9600-0.9789). Across the smoothing alternatives and evaluated outputs, the median ratio of alternative to reference valid-voxel counts ranged from 99.953% to 100%. This count ratio describes coverage, not equality of voxel locations. Aggregate sensitivity results are provided in the linked analysis supplement.

Timing quantities were more threshold-dependent. Across DTP, DTR, refined total lag and relative arrival, median spatial r ranged from 0.7019 to 0.9158. For 5%/95%, DTP1 and both DTR intervals typically lengthened by 1 TR. For 20%/80%, DTP1 shortened by 2 TR and both DTR intervals by 1 TR. These changes are expected consequences of changing the operational transition thresholds. Timing maps were also more sensitive to smoothing than Hypoxia_SS_: the lowest median correlations were 0.627 for DTR1 with the 8-volume temporal window and 0.557 for DTP2 with 4-mm spatial smoothing. The sensitivity analysis therefore supports robustness of the steady state pattern within the tested range, but not parameter invariance of the timing estimates or patient classifications.

### 3.3 Iteration and Termination Behavior

Across 1,827,042 mask voxels, the pooled refinement-count median and interquartile range were 2 [2, 2] for all four estimators. The numerical criterion was satisfied in 97.789% of evaluable DTP1 estimates and 97.682% of DTP2 estimates. 2.211% and 2.318%, respectively, remained criterion-unsatisfied at bounded termination. All evaluable DTR1 and DTR2 estimates satisfied their criterion. Maximum refinement counts were 17, 17, 9 and 11 for DTP1, DTP2, DTR1 and DTR2. A crossing fallback was encountered at least once in 20.028%, 1.168%, 0.006% and 0.002% of mask voxels, respectively. An encounter does not prove that the final retained index was itself a fallback. DTP2 recorded 538 caught voxel errors and 609 missing outputs. DTR2 also had 609 missing outputs. These findings describe numerical behavior, not physiological equilibration.

### 3.4 G-adjusted Normative Model

The validated G-adjusted model had a median in-sample voxel-wise R^2^ of 0.490. In leave-one-subject-out validation, the median voxel-wise predictive Q^2^ was 0.345 across 329,449 eligible voxels. Compared with the training-mean model, G adjustment reduced pooled RMSE from 0.02195 to 0.01843 (16.0%) and improved subject-level RMSE in 19 of 20 controls. In-sample R^2^ and cross-validated Q^2^ describe different estimands and are reported separately. G adjustment was used for the proof-of-concept Hypoxia_SS_ patient maps. DTP, DTR and Delay_corr_ used conventional atlas z-scores.

### 3.5 Proof-of-Concept Patient Maps

Representative patient maps are shown in Figures 2 and 3. Across the three cases, the timing, steady state and deviation maps displayed spatially heterogeneous patterns within and around the structurally defined tumor environment. The side-by-side whole-stimulus map provided visual context for how temporal averaging can reduce the conspicuity of a localized steady state pattern. This was most apparent in the third case (Figure 3), in which the steady state map and its deviation map showed a segmental spatial pattern that was less conspicuous in the whole-stimulus summary. The cases are illustrative only: no patient-level hypothesis testing was performed, and the visual patterns do not establish tumor specificity, diagnostic accuracy, or biological truth.

**Fig. 2:**
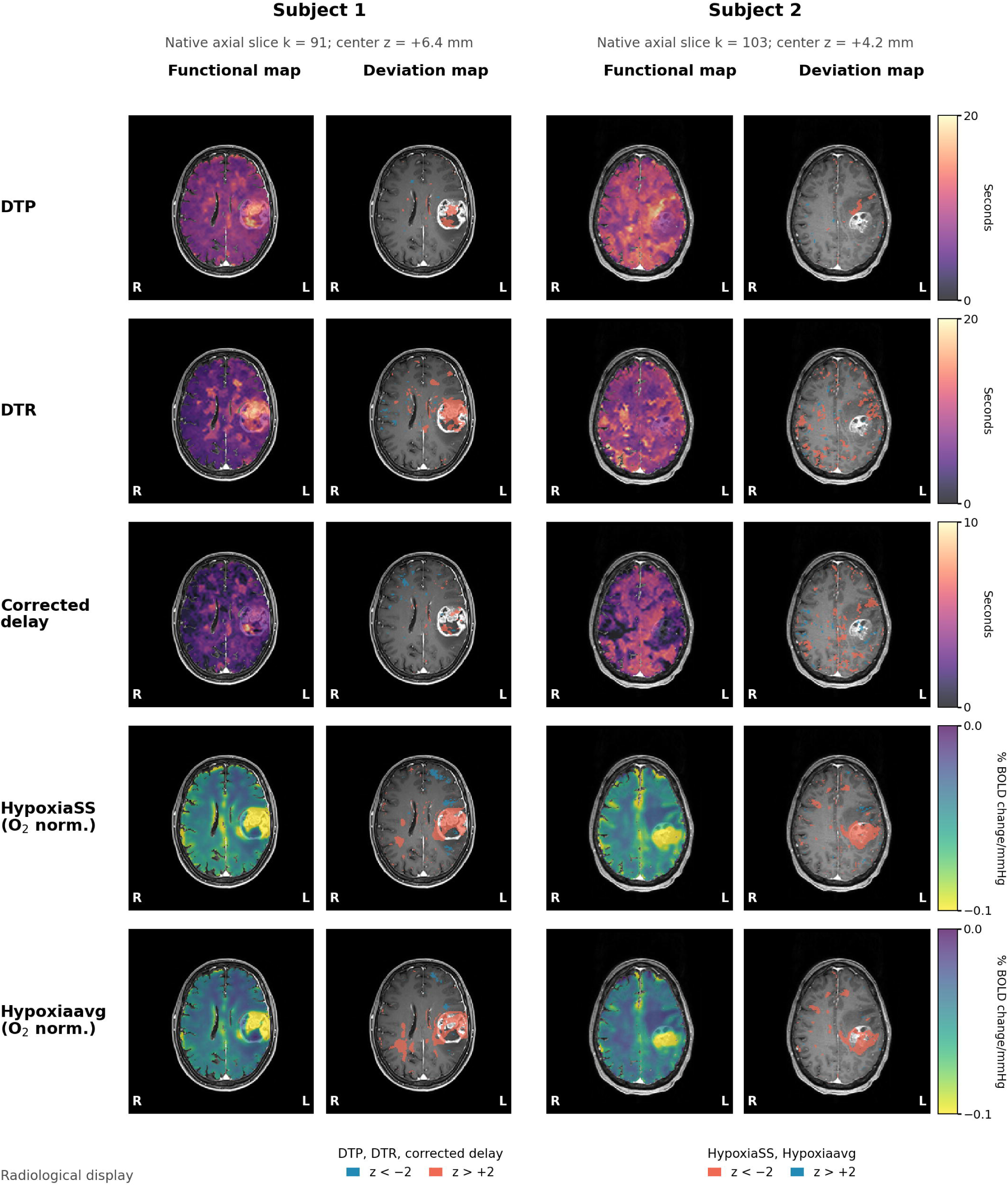
Parametric and deviation maps for two representative patients with newly diagnosed glioblastoma. Subject 1 is shown at native axial slice k = 91 (center z = +6.4 mm). Subject 2 is shown at native axial slice k = 103 (center z = +4.2 mm). Each row corresponds to DTP, DTR, relative arrival delay (Delay_corr_), Hypoxia_SS_ O_2_-normalized, and Hypoxia_avg_ O_2_-normalized. For each subject, the functional map is paired with the corresponding deviation map. Timing maps use magma (DTP and DTR 0-20 s, Delay_corr_ 0-10 s), and response maps use reversed viridis (−0.10 to 0 %BOLD/mmHg). For timing deviation maps, z < −2 is blue and z > +2 is red. For response deviation maps, z < −2 is red and z > +2 is blue. Hypoxia_SS_ deviations use G-adjusted standardized residuals. All other deviations use conventional atlas z-scores. The images are shown in radiological orientation.

**Fig. 3:**
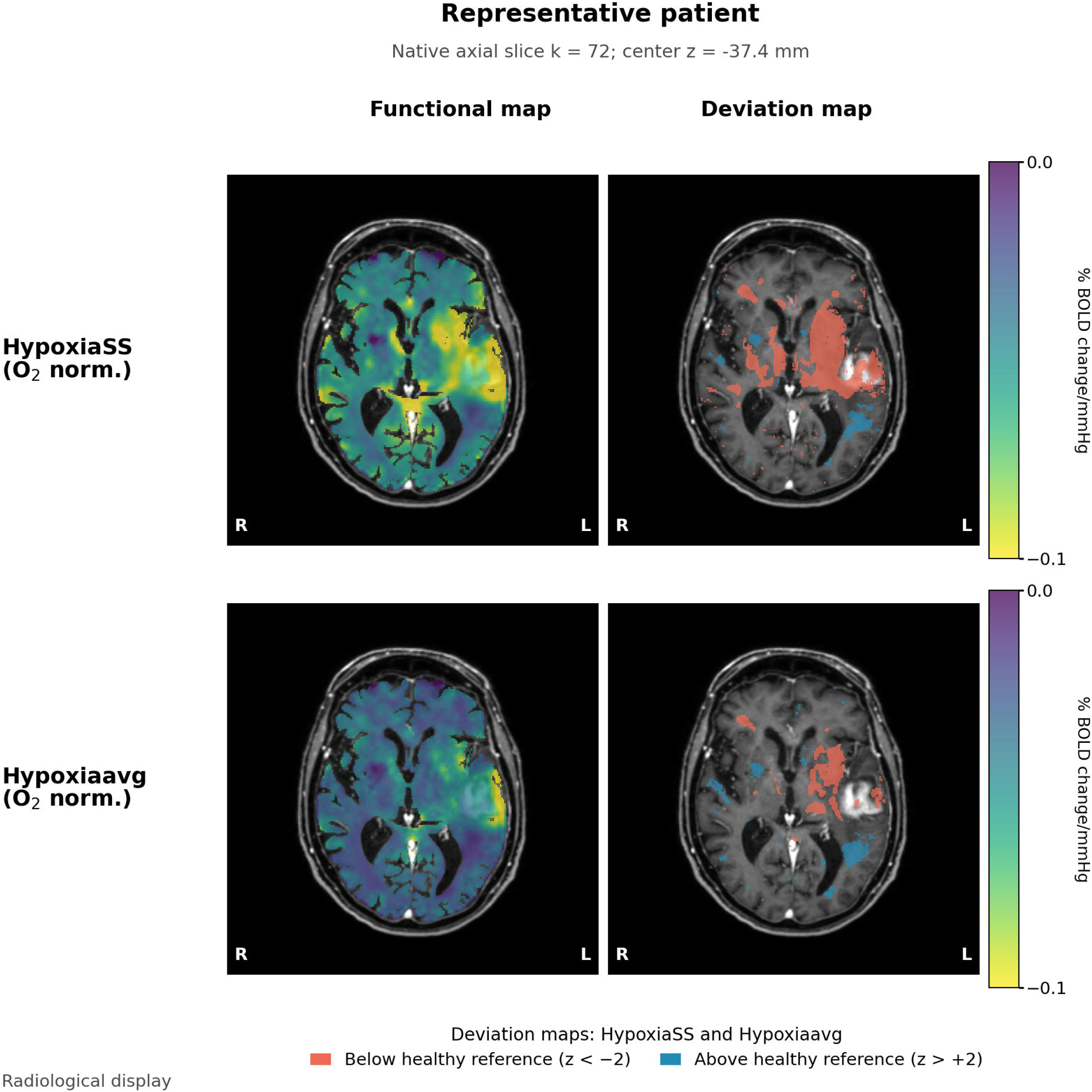
Functional and deviation maps for Hypoxia_SS_ O_2_-normalized and Hypoxia_avg_ O_2_-normalized in a representative patient with glioblastoma at native axial slice k = 72 (center z = −37.4 mm). Functional maps use reversed viridis from −0.10 to 0 %BOLD/mmHg. In the deviation maps, below-reference values (z < −2) are red and above-reference values (z > +2) are blue. Hypoxia_SS_ deviations use G-adjusted standardized residuals. Hypoxia_avg_ deviations use conventional atlas z-scores. The images are shown in radiological orientation. The side-by-side display illustrates how temporal summaries may differ spatially and is not an accuracy comparison.

## 4 Discussion

### 4.1 Methodological Contribution

GLIODISSECT separates a voxel’s response into relative arrival, descending transition, threshold-defined steady state, and recovery transition. These outputs are mathematically related, but they answer different descriptive questions: when the response arrives, how long the transitions last, and what response magnitude remains after the descending transition. In a heterogeneous tumor environment, this separation offers a more interpretable representation than assigning the entire time course a single summary value. Voxel-wise landmark refinement is central to this approach because it adapts the analysis windows to the local response. The steady state amplitude can therefore be interpreted alongside the time required for the response to develop and recover. This is particularly relevant when neighboring voxels within the same structural tumor compartment exhibit different response dynamics. The component maps preserve their spatial correspondence, allowing these differences to be examined together and expressed as deviations from their respective healthy references. GLIODISSECT thus links the temporal organization of the BOLD response to anatomically resolved mapping of functional heterogeneity.

### 4.2 Physiological Interpretation

The physiological value of the steady state map is that it quantifies the BOLD response within each voxel’s threshold-defined plateau, separating steady state amplitude from arrival delay and transition kinetics. Controlled hypoxia increases deoxyhemoglobin and generates oxygenation-sensitive BOLD contrast [14, 19]. For a given acquisition and stimulus, response magnitude depends on the vascular volume contributing to the signal and the change in blood oxygenation [14]. The temporal maps provide complementary information about the development and recovery of this response. In healthy volunteers, Harris et al. observed that oxygenation-sensitive R_2_* changes preceded the slower cerebral blood-flow response during a 20-minute isocapnic hypoxic challenge [20]. This finding supports considering oxygenation and flow dynamics separately. Stimulus duration, severity and gas control must be taken into account in comparisons across studies [14, 20].

In tumor tissue, prolonged DTP and altered Delay_corr_ may reflect dispersion of the hypoxic response through irregular vascular pathways, functional shunting and heterogeneous microvascular transit [21, 22]. A voxel with delayed arrival or a prolonged transition can still reach a pronounced steady state response. Examining these maps together therefore distinguishes temporal abnormalities from differences in steady state amplitude. The threshold-based iterative approach identifies defined fractions of each voxel’s signal excursion and does not require a fixed parametric response shape. The specific contributions of vascular volume, oxygen extraction and tumor infiltration require assessment with spatially matched perfusion imaging and histopathology.

### 4.3 Normative and Patient-specific Mapping

The normative atlas converts the component maps from descriptive images into voxel-wise deviations from a healthy reference. Voxel-wise comparison with a healthy reference has precedent in CVR imaging [23]. The cross-validated 16.0% pooled RMSE reduction shows that the global response scalar G contains useful out-of-sample information for predicting healthy-control Hypoxia_SS_, while the median Q^2^ of 0.345 also shows that substantial local variation remains unexplained. Because G is derived from Hypoxia_SS_ itself, it is an internal adjustment rather than an independent physiological predictor. In the patient cases, the component-specific z maps made spatial heterogeneity visible relative to the healthy reference. This is the intended proof of concept: the framework supports anatomically resolved interpretation of different temporal features. It does not establish whether a deviation represents tumor infiltration or any specific vascular or metabolic mechanism.

### 4.4 Limitations

The healthy reference cohort was modest and young, and the three patient cases were descriptive. Age- and sex-related effects could not be modeled. The G-adjusted model improved held-out prediction but remained a single-covariate internal adjustment. The patient tumor-excluded G mask differs from the control mask and may affect deviation estimates. Very small positive warped normative standard deviations produced extreme unclipped z-score tails. Those tails should not be interpreted as biological severity. Threshold perturbations preserved steady state map patterns, but timing estimates were more dependent on the operational thresholds and smoothing choices. DTP1 crossing fallbacks were common despite high overall criterion satisfaction. The full-record lag initializer and established search windows do not guarantee sample-level independence of the first step from later time points. The whole-stimulus map is not an established benchmark for this paradigm, and visual differences between maps do not establish superiority. No independent biological reference was available. The numerical audit did not establish anatomically specific regions in which estimator performance was better or worse. Future work should externally validate the model in larger age-matched cohorts, assess separate-visit repeatability, evaluate additional physiological and technical covariates, and include spatially matched histopathological, perfusion, molecular-imaging, or longitudinal validation. Potential acquisition improvements include shorter-TR or multiband EPI, multi-echo acquisition, and optimized receiver coils. These approaches and any increase in spatial resolution would require balancing temporal sampling, signal-to-noise ratio and partial-volume effects.

## 5 Conclusion

GLIODISSECT provides operational voxel-wise maps of relative arrival, transition timing and steady state amplitude during controlled hypoxia. The technical analyses support numerical stability of most estimates, steady state map-pattern robustness within the tested threshold and smoothing ranges, and useful cross-validated global-response adjustment. In three proof-of-concept glioblastoma cases, component-specific maps and normative deviations visualized spatially heterogeneous response features that cannot be represented by one summary value. These findings establish a method for interpretable temporal and voxel-wise mapping, not diagnostic superiority or biological validation.

## Data Availability Statement

Group-level healthy-control hypoxia-powered BOLD MRI reference and normative-model maps are available from Mendeley Data at https://doi.org/10.17632/2k4xh888b4.1. Patient imaging data cannot be deposited publicly because of institutional data-governance requirements and ethical restrictions. GLIODISSECT, the MATLAB temporal-decomposition software, is available at https://github.com/tristanschmidlechner-oss/GLIODISSECT and archived on Zenodo at https://doi.org/10.5281/zenodo.23024999. The separate manuscript-analysis supplement is available at https://github.com/tristanschmidlechner-oss/GLIODISSECT-analysis. It provides normative-model and leave-one-out cross-validation code, threshold and smoothing sensitivity workflows, convergence instrumentation, synthetic examples and aggregate reference results. The supplement requires GLIODISSECT and compatible prepared inputs. Participant-level study data and a complete registration or figure-generation workflow are not included.

## CRediT Author Statement

Tristan Schmidlechner: Conceptualization, Data curation, Formal analysis, Methodology, Software, Visualization, Writing – original draft, Writing – review & editing. Vittorio Stumpo: Investigation, Writing – review & editing. Elisabeth Jehli: Investigation, Writing – review & editing. Leonie Zerweck: Investigation, Writing – review & editing. Jacopo Bellomo: Investigation, Writing – review & editing. Meltem Gönel: Investigation, Writing – review & editing. Flavia Müller: Investigation, Writing – review & editing. Natalia Cantavella Franch: Investigation, Writing – review & editing. Martina Sebök: Investigation, Writing – review & editing. Andrea Bink: Investigation, Resources, Writing – review & editing. Zsolt Kulcsár: Investigation, Resources, Writing – review & editing. Michael Weller: Resources, Supervision, Writing – review & editing. Luca Regli: Resources, Supervision, Writing – review & editing. Jorn Fierstra: Project administration, Supervision, Writing – review & editing. Christiaan H. B. van Niftrik: Conceptualization, Project administration, Supervision, Writing –review & editing.

## Declaration of Generative AI and AI-Assisted Technologies in the Manuscript Preparation Process

During preparation of this work, the authors used Claude (Anthropic) and ChatGPT/Codex (OpenAI) to assist with code review, analysis scripting, manuscript structuring and editorial revision. The authors reviewed and validated all analyses, code outputs and text, made all scientific decisions, and take full responsibility for the published article.

